# Direct Comparison of SARS-CoV-2 Nasal RT-PCR and Rapid Antigen Test (BinaxNOW™) at a Community Testing Site During an Omicron Surge

**DOI:** 10.1101/2022.01.08.22268954

**Authors:** John Schrom, Carina Marquez, Genay Pilarowski, Grace Wang, Anthea Mitchell, Robert Puccinelli, Doug Black, Susana Rojas, Salustiano Ribeiro, Jacqueline Martinez, Diane Jones, Robert Nakamura, Vivek Jain, Maya Petersen, Joe DeRisi, Diane Havlir

## Abstract

In 731 persons seeking COVID-19 testing at a walk-up San Francisco community site in January 2022, simultaneous nasal rapid antigen testing (BinaxNOW™) and RT-PCR testing was performed. There were 296 (40.5%) positive tests by RT-PCR; 98.5% of a random sample (N=67) were the omicron variant. Sensitivity of a single antigen test was 95.2% (95% CI 92-98%); 82.1% (95% CI 77-87%) and 65.2% (95% CI 60-71%) for Ct threshold of < 30, < 35 and no threshold, respectively. We also compared BinaxNOW™ to RT-PCR from oral cheek swabs to nasal swabs (N=75); oral cheek specimen was significantly less sensitive than nasal swab. A single BinaxNOW™ oral cheek rapid antigen test failed to detect 91% (20 of 22) of specimens that were BinaxNOW™ positive from the standard nasal sampling. In a separate direct comparison of BinaxNOW™ between specimens collected from nasal or throat (tonsillar) swab (N=115), sensitivity was 97.7% for nasal and 48.8% for throat swabs that were PCR-positive on nasal swab with a Ct threshold < 30. Among persons with either a nasal or throat RT-PCR positive swab with Ct<30, BinaxNOW™ sensitivity was 85.7% for nasal and 89.8% for nasal plus throat swabs. BinaxNOW continues to be a very useful diagnostic during the omicron surge; oral (throat or cheek swab) should not replace nasal swabs due to significantly reduced sensitivity compared to nasal. As currently recommended, repeat testing should be done for high-risk persons with an initial negative antigen test result.

## Introduction

SARS-CoV-2 rapid antigen tests are a valuable public health tool in the ongoing COVID-19 pandemic. They enable immediate identification of active SARS-CoV-2 infection with high viral levels, which can lead to faster isolation and curtail transmission chains (1). Antigen tests also enable rapid diagnosis needed for initiation of time-sensitive outpatient COVID-19 therapies. Many community testing programs, including school programs, utilize rapid tests; home antigen testing is increasingly becoming available in the United States and is widely available in some countries. Widespread use of rapid antigen tests is based on performance evaluations in the context of the ancestral, alpha, or delta variants (2).

It is vital to evaluate performance of rapid antigen tests when new SARS-CoV-2 variants emerge. The omicron variant has over 50 mutations compared to ancestral lineages; the majority are in the spike protein, but 4 are in the nucleocapsid gene, the target gene in the BinaxNOW assay. While laboratory studies suggest that the performance of BinaxNOW should not be affected when used to detect the omicron variant, field validation is needed (3). We sought to examine performance of the BinaxNOW rapid antigen tests at our community-based site in the Mission District in San Francisco, a setting in which we have routinely used this test and have evaluated its performance with previous variants (4,5).

## Methods

This report includes data collected January 3-4, 9^th^ and 14th 2022 at our free, outdoor, walk-up testing and vaccine site situated in an outdoor parking lot in the heart of the Mission Cultural District in San Francisco. The site is led by Unidos en Salud – an academic (UCSF and Chan Zuckerberg Biohub), community (Latino Task Force), and public health (San Francisco Department of Public Health) collaboration that conducts SARS-CoV-2 surveillance and serves communities at highest COVID-19 risk (4,5,6). Unidos en Salud serves a San Francisco community with a high proportion of frontline workers, immigrants, and monolingual families living in multi-generational households, the majority being Latinx.

Persons seeking testing provided demographics, symptoms and onset date, vaccination status, reason for testing and informed consent. There was no age restriction. Certified lab assistants collected bilateral anterior nasal swabs using manufacturer instructions. A second anterior nasal swab was immediately collected for RT-PCR. Certified readers read BinaxNOW cards, and results were returned within an hour of testing using secure messaging in the Primary.Health platform. Photographed cards were re-read by a blinded trained expert, and these results were considered final for analyses. On January 9, we collected simultaneously anterior nasal swabs and oral cheek swabs on 75 persons randomly selected from persons seeking testing that day and tested for SARS CoV-2 with BinaxNOW and RT-PCR. On January 14, we collected simultaneously anterior nasal swabs and oral tonsillar swabs on 115 persons randomly selected from persons seeking testing that day and tested for SARS CoV-2 with BinaxNOW and RT-PCR. Bilingual (Spanish and English) Unidos en Salud staff called persons diagnosed with COVID-19, offering supportive services including home deliveries of supplies, food, and care items as previously described (6). Persons who were eligible for COVID-19 treatment were referred to their primary health provider or Zuckerberg San Francisco General Hospital. Persons at risk with negative test results were advised to seek repeat rapid and/or RT-PCR testing.

RT-PCR using probes specific to SARS-CoV-2 N and E genes were performed on the nasal swabs collected in DNA/RNA Shield (Zymo Research) with an internal human positive control (RNase P) as previously described (7). The assay limit of detection is 100 viral copies/mL; cycle thresholds less than 40 were considered positive. Using the same isolated RNA, allele specific RT-PCR was used to discriminate the two dominate lineages, omicron and delta, according to the manufacturer’s directions (8).

We calculated sensitivity and specificity of the rapid antigen tests, with RT-PCR as gold standard, both overall and using RT-PCR cycle thresholds (Ct) below 30 and 35 with the 731 specimens collected on January 3,4. 95% confidence intervals were calculated using the Clopper-Pearson method. We examined assay performance in strata defined by presence versus absence of symptoms at time of testing, age and vaccination status. We also calculated sensitivity and specificity of the rapid antigen tests, with RT-PCR as the gold standard, both overall and using RT-PCR cycle thresholds (Ct) below 30 and 35 with the 75 nasal and oral cheek specimens collected on January 9. We did similar analyses for the 115 persons providing nasal and oral tonsillar specimens collected on January 14.

### Ethics Statement

The UCSF Committee on Human Research determined that the study met criteria for public health surveillance. All participants provided informed consent for dual testing.

## Results

SARS-CoV-2 RT-PCR and BinaxNOW testing was performed on 731 samples over a two-day period on January 3 and 4 at our community testing site. Participants self-identified as 77.1% Latinx, 6.9% American, Central or South American Indian, 5.1% white, 3.2% Asian and 1.8% Black. There were 52.6% males and 47.4% females; 15.3% were under 12 years of age. Participants reported that the primary motivation for testing was clearance for work (26.1%), known or suspected exposure (23.1%), school requirement (17.8%), and due to symptoms (16.7%).

Overall, 296 of 731 (40.5%) persons tested were SARS CoV-2 positive by RT-PCR. In a convenience? sample of 184 samples, 66 samples, all with a Ct threshold of < 35, were consistent for omicron by allele-specific PCR using (8), and 1 sample was consistent for the delta variant. Similar to performance with other variants, the BinaxNOW assay had its highest performance among persons with low Ct values. There were 177 persons BinaxNOW positive among the 186 persons RT-PCR positive with Ct <30. There were no BinaxNOW positives among the 61 persons RT-PCR positive with Ct >35. (Figure 1). We calculated sensitivity of BinaxNOW within Ct threshold of < 30, < 35 and overall (i.e., no threshold) as 95.2% (95% CI 92-98%); 82.1% (95% CI 77-87%) and 65.2% (95% CI 60-71%), respectively (Table 1). BinaxNOW performance was similar among persons less than and greater than 12 years of age.

**Table 1:**
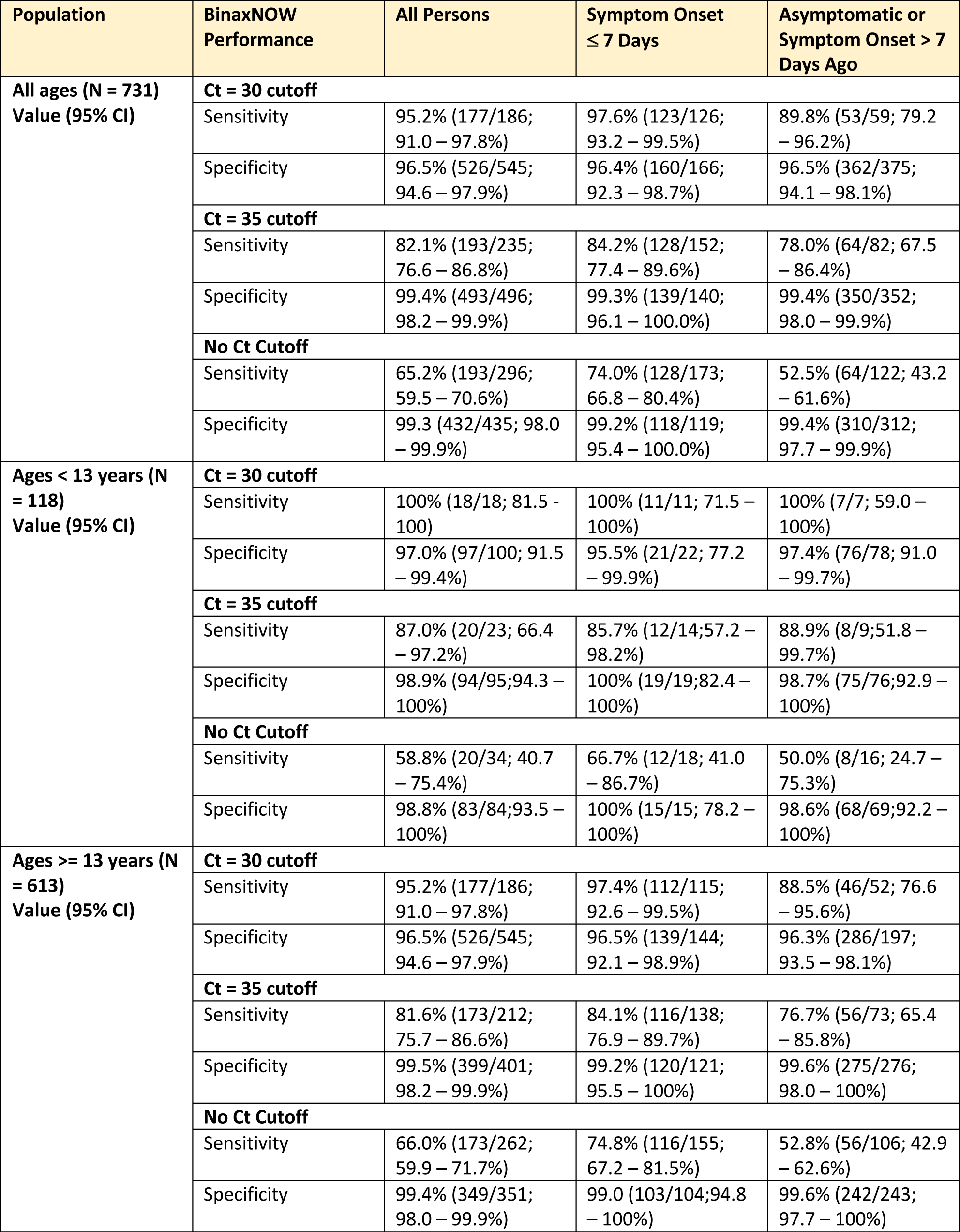
Sensitivity and Specificity of BinaxNOW Stratified by Age and Symptoms.

**Figure 1.**
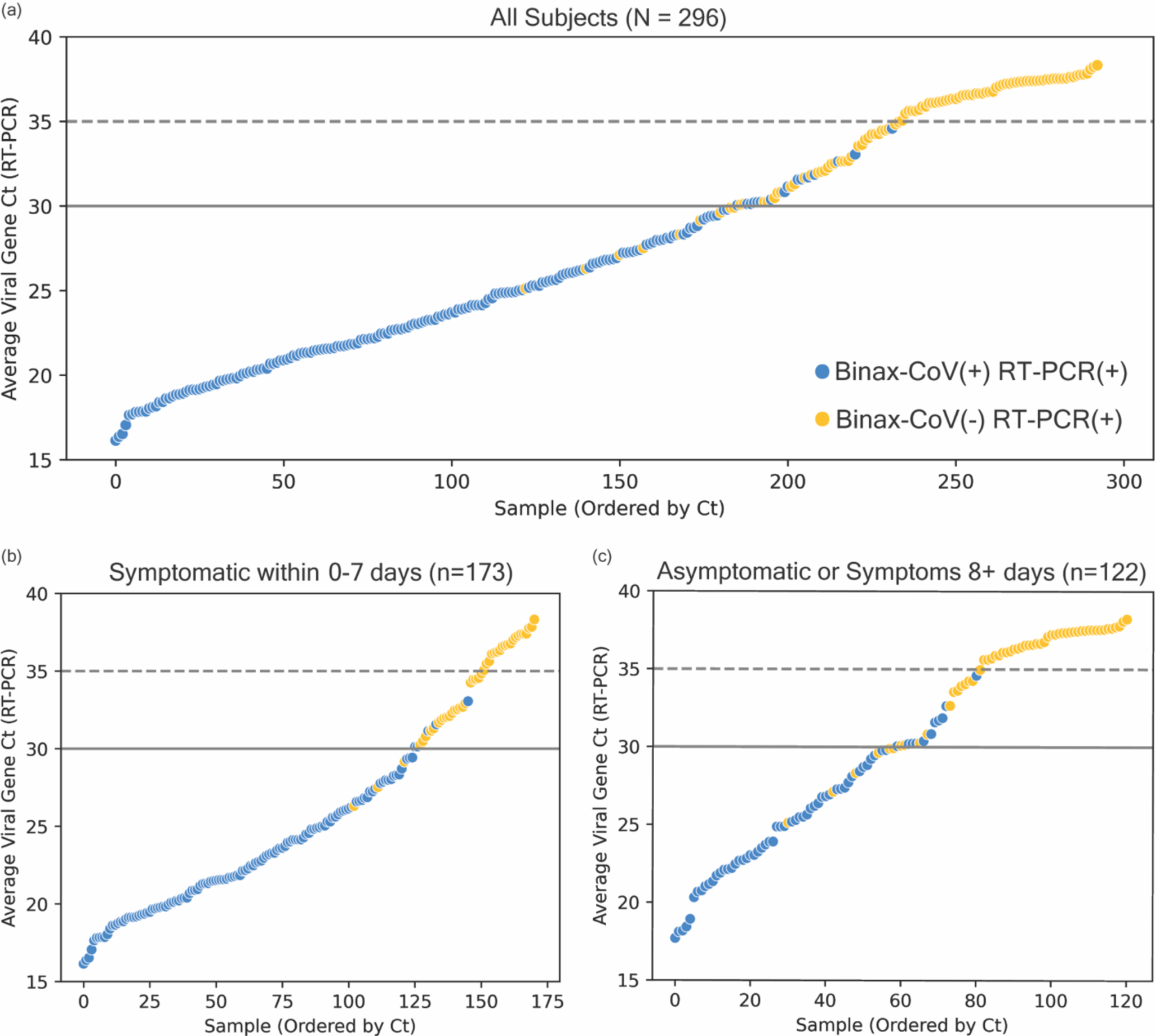
RT-PCR Ct values and BinaxNOW rapid antigen test results of all participants tested January 3 and 4 (a), and stratified according to COVID-19 symptoms (b and c). Average viral Ct values of all individuals with positive RT-PCR and/or rapid antigen test results (N = 296 total) are plotted in ascending order of Ct. Each point represents one individual. Blue circles are individuals whose samples were positive on both rapid antigen test (BinaxNOW) and on RT-PCR test. Yellow circles represent individuals who were RT-PCR positive, but rapid antigen test negative. Abbreviations: COVID-19, coronavirus disease 2019; Ct, cycle threshold; RT-PCR, reverse transcription polymerase chain reaction.

When stratified by symptom status, the BinaxNOW detected 53 of 59 RT-PCR positive persons who were asymptomatic for a sensitivity of 89.8% (95% CI 79-96%); sensitivity was 97.6% (95% CI 93-100%) for persons with symptoms. (Figure 1 and Table 1). We note that the BinaxNOW sensitivity was slightly lower but overlapping across strata of vaccination (Table 2). Specificity remained greater than 95% for all strata and Ct thresholds considered.

**Table 2:**
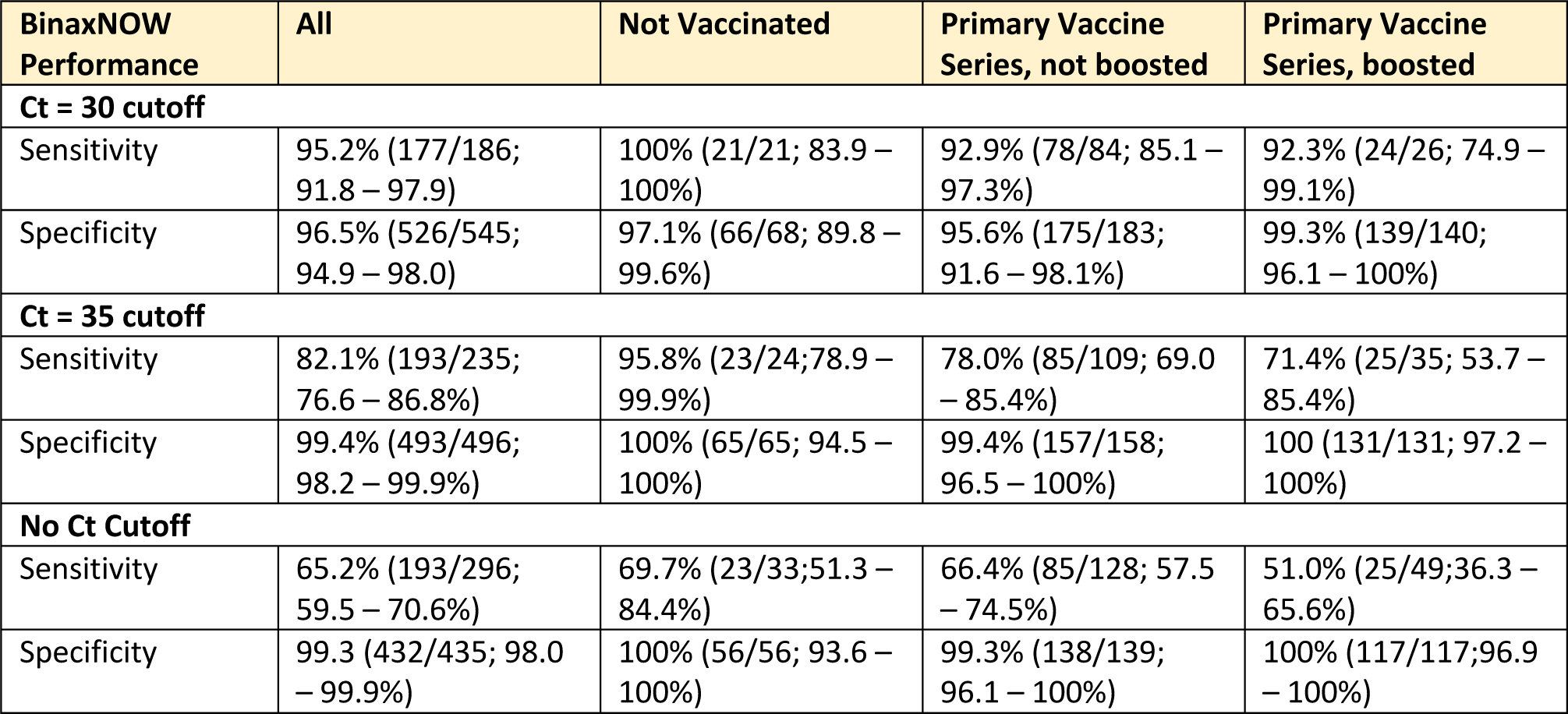
Sensitivity and Specificity of BinaxNOW Stratified by Vaccination Status.

Of the simultaneous nasal and oral cheek specimens collected on 75 persons on January 9, 46/75 (61%) were positive on RT-PCR from the nares (Table 3),with 22 specimens yielding a Ct of less than 30. Among the 46 nasal RT-PCR positives, 22 were positive by BinaxNOW, while only 2 were positive from oral cheek specimens with BinaxNOW (4.3% sensitivity). Thirteen of the 46 RT positive nasal specimens were also positive on oral cheek RT-PCR. The sensitivity of the oral cheek specimens was only 9.0% for Ct threshold <30. There were no specimens RT-PCR positive from the oral cheek collection and negative on the nasal RT-PCR.

**Table 3.** Sensitivity and Specificity for BinaxNOW from Nasal or Oral Cheek Specimen Collection from 75 specimens collected.

| BinaxNOW Performance | Nasal Swab<sup>1</sup> | Cheek Swab<sup>1</sup> | Cheek Swab<sup>2</sup> |
| --- | --- | --- | --- |
| **Ct = 30 cutoff** |  |  |  |
| Sensitivity | 86.4% (19/22; 65.1 – 97.1%) | 9.1% (2/22; 1.1 – 29.2%) | 100% (1/1; 3 – 100%) |
| Specificity | 94.3% (50/53; 84.3 – 98.8%) | 100% (53/53; 93.3 – 100%) | 98.6% (73/74; 92.7 – 100%) |
| **Ct = 35 cutoff** |  |  |  |
| Sensitivity | 74.1% (20/27; 53.7 – 88.9%) | 7.4% (2/27; 0.9 – 24.3%) | 50% (1/2; 2.5 – 98.7%) |
| Specificity | 95.8% (46/48; 85.7 – 99.5%) | 100% (48/48; 92.6 – 100%) | 98.6% (72/73; 92.6 – 100%) |
| **No Ct Cutoff** |  |  |  |
| Sensitivity | 47.8% (22/46; 32.9 – 63.1%) | 4.3% (2/46; 0.5 – 14.8%) | 7.7% (1/13; 1.9 – 36.0%) |
| Specificity | 100% (29/29; 88.1 – 100%) | 100% (29/29; 88.1 – 100%) | 98.4% (61/62; 91.3 – 100%) |

<sup>1</sup>Referent (denominator) is positive on RT-PCR from nasal specimen
<sup>2</sup>Referent (denominator) is positive on RT-PCR from oral cheek specimen

In the comparison between nasal and oral tonsillar specimens obtained on 115 persons on January 14, the sensitivity for detecting PT-PCR positive cases with Ct <30 and Ct<30-35 was 97.7% and 82.0%, respectively, using the nasal swab alone, and 74.1% and 54.5% using the oral throat swab alone (Table 4). For persons with a PCR positive swab (Ct <30) from either nasal or throat specimen, the sensitivity of BinaxNOW was 85.7% for nose alone, 46.9% for throat alone and 89.8% for either nose or throat (Figure 2).

**Table 4.** Sensitivity and Specificity for BinaxNOW from Nasal or Throat Cheek Specimen Collection from 115 specimens collected.

| **BinaxNOW**<br>**Performance** | **Nasal BinaxNOW**<br>**Referent Nasal**<br>**RT-PCR** | **Throat**<br>**BinaxNOW**<br>**Referent Throat**<br>**RT-PCR** | **Throat**<br>**BinaxNOW**<br>**Referent Nasal**<br>**RT-PCR** | **Nasal BinaxNOW**<br>**Referent: Either**<br>**nasal or throat**<br>**RT-PCR** | **Throat**<br>**BinaxNOW**<br>**Referent: Either**<br>**nasal or throat**<br>**RT-PCR** | **Either Nasal or**<br>**Throat**<br>**BinaxNOW**<br>**Referent: Either**<br>**nasal or throat** |
| --- | --- | --- | --- | --- | --- | --- |
| **Ct = 30 cutoff** |  |  |  |  |  |  |
| Sensitivity | 97.7% (42/43;<br>87.7 – 99.9%) | 74.1% (20/27;<br>53.7 – 88.9%) | 48.8% (21/43;<br>33.3 – 64.5%) | 85.7% (42/49;<br>72.8 – 94.1%) | 46.9% (23/49;<br>32.5 – 61.7%) | 89.8% (44/49;<br>77.8 – 96.6%) |
| Specificity | 83.3% (60/72;<br>72.7 – 91.1%) | 92.0% (81/88;<br>84.3 – 96.7%) | 91.7% (66/72;<br>82.7 – 96.9%) | 81.8% (54/66;<br>70.4 – 90.2%) | 93.9% (62/66;<br>85.2 – 98.3%) | 77.3% (51/66;<br>65.3 – 86.7%) |
| **Ct = 35 cutoff** |  |  |  |  |  |  |
| Sensitivity | 82.0% (50/61;<br>70.0 – 90.6%) | 54.5% (24/44;<br>38.8 – 69.6%) | 39.3% (24/61;<br>27.1 – 52.7%) | 73.5% (50/68;<br>61.4 – 83.5%) | 36.8% (25/68;<br>25.4 – 49.3%) | 77.9% (53/68;<br>66.2 – 87.1%) |
| Specificity | 92.6% (50/54;<br>82.1 – 97.9%) | 95.7% (68/71;<br>88.1 – 99.1%) | 94.4% (51/54;<br>84.6 – 98.8%) | 91.5% (43/47;<br>79.6 – 97.6%) | 95.7% (45/47;<br>85.5 – 99.5%) | 87.2% (41/47;<br>74.3 – 95.2%) |
| **No Ct Cutoff** |  |  |  |  |  |  |
| Sensitivity | 56.5% (52/92;<br>45.8 – 66.8%) | 44.8% (26/58;<br>31.7 – 58.5%) | 28.3% (26/92;<br>19.4 – 38.6%) | 54.2% (52/96;<br>43.7 – 64.4%) | 28.1% (27/96;<br>19.4 – 38.2%) | 59.4% (57/96;<br>48.9 – 69.3%) |
| Specificity | 91.3% (21/23;<br>72.0 – 98.9%) | 98.2% (56/57;<br>90.6 – 100%) | 95.7% (22/23;<br>78.1 – 99.9%) | 89.5% (17/19;<br>66.9 – 98.7%) | 100% (19/19;<br>82.4 – 100%) | 89.5% (17/19;<br>66.9 – 98.7%) |

**Figure 2.**
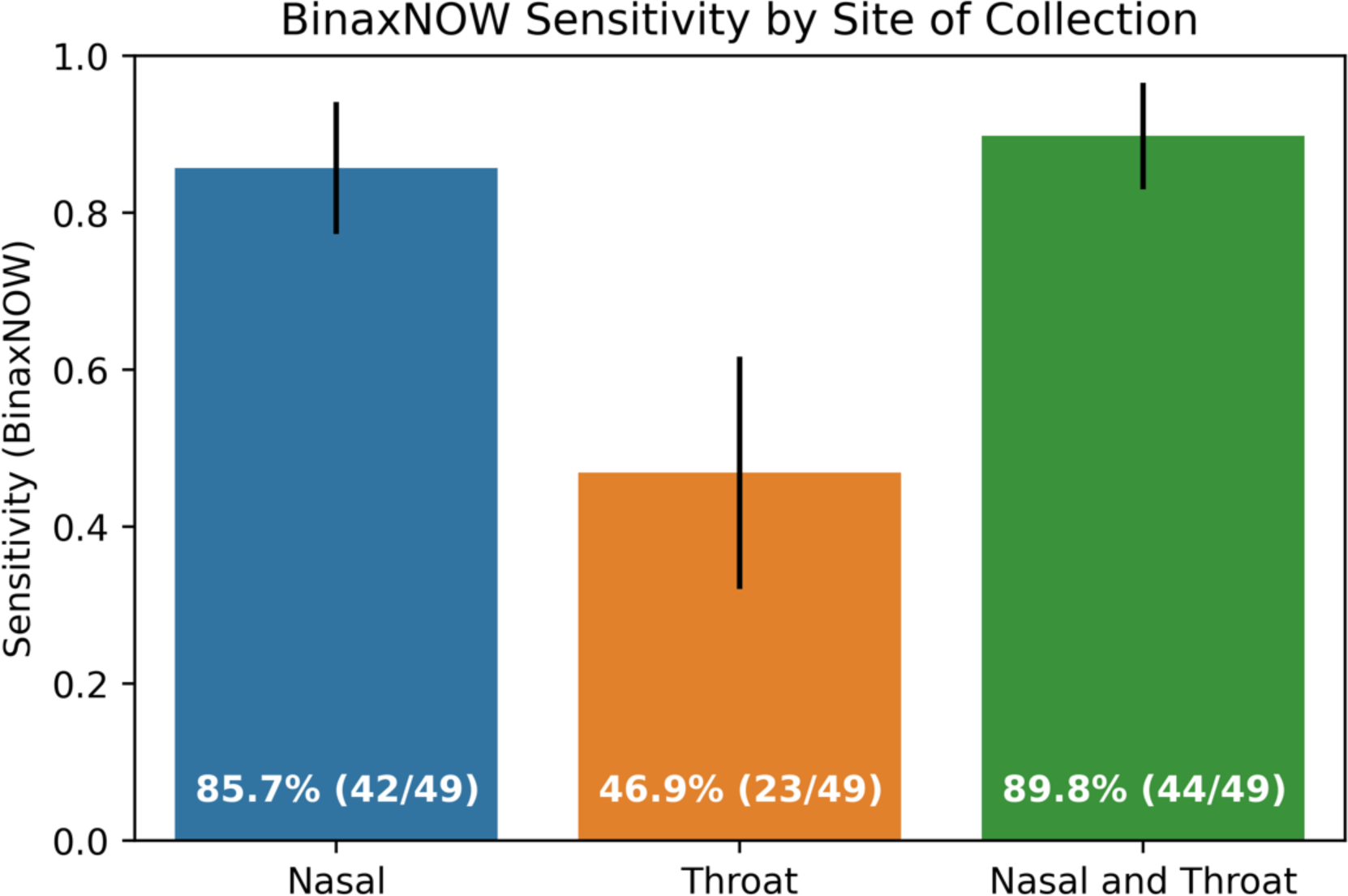
Sensitivity of BinaxNOW tests by site of collection based on samples collected on January 14 (N=115). Sensitivity is calculated using either nasal or throat RT-PCR as the referent for samples with Ct less than 30. Bar height indicates the calculated sensitivity for the indicated site of collection; error bars indicate the 95th percentile confidence interval as calculated using the Clopper-Pearson method. ‘Nasal and throat’ indicates a BinaxNOW positive result in either the nasal or throat or both samples. The calculated sensitivity, expressed both as a percent and a fraction, is overlaid on the corresponding bar. Abbreviations: COVID-19, coronavirus disease 2019; Ct, cycle threshold; RT-PCR, reverse transcription polymerase chain reaction.

## Discussion

This cross-sectional analysis confirms that the BinaxNOW rapid antigen test detects omicron with a sensitivity similar to that observed for prior variants. The assay rapidly identifies persons with highest levels of virus, and thus those likely to pose the greatest risk for transmission at the time of the test (9). A positive rapid test enables immediate public health and personal action for isolation, disease mitigation, and clinical care, in a disease process where chains of transmission need to be broken and therapies are time-sensitive. With the increasing availability of this test in the United States, this information can inform optimal emerging public health strategies that hinge on rapid diagnosis and treatment.

Our data support the recommendation for repeat rapid antigen testing for persons at risk for COVID-19 who have an initial negative BinaxNOW result. Persons who have low levels of virus detectable on PCR but not antigen test may be either at the upswing or downswing of the viral dynamic curve for SARS Co-V-2 (10). In the setting of an acute surge, it is likely than many persons are on the upswing, and may subsequently develop higher viral loads associated with greater infectiousness and detectable on repeat testing 1-2 days later.

Importantly, our data are cross sectional and cannot inform questions on natural history of infection. Omicron putatively has a shorter incubation period than prior variants. Symptoms are more intense in the upper respiratory track, and the virus appears to be less pathogenic in the lower airway compared to prior variants (11). One small study with five positive health care workers suggested detection is faster from oral versus nasal swabs (12). Although we only examined 75 paired nasal and oral cheek swabs in our cross-sectional analyses, our data are compelling that a simple oral cheek swab does not increase detection of SARS-CoV-2. Further, the throat swab significantly underperformed compared to the standard nasal swab. Even with the prominent clinical feature of pharyngitis in persons with omicron, our data argue against replacing nasal swabs with throat swabs for diagnosis. We saw only a small (< 5% increase) in detection of COVID by adding a throat to a nasal swab. In a high throughput community testing site, the complexity and cost of such an approach would need to be formally evaluated before adopting.

## Data Availability

All data produced in the present study are available upon reasonable request to the authors

